# Determinants of obesity and overweight among school-aged children in Bauchi, Nigeria: A mixed-methods study

**DOI:** 10.1101/2025.08.28.25334654

**Authors:** Umar Ahmad, Nazif Yakubu, Grace Joseph, Murtala Muhammad Jibril, Aminu Umar Kura

**Author notes:** Corresponding author: Dr. Umar Ahmad.

## Abstract

**Background:** Childhood obesity is an escalating public health issue with serious long-term implications. This study investigates the factors contributing to overweight and obesity among school-aged children in Bauchi, Nigeria, utilizing a mixed-methods design.

**Materials and methods:** A cross-sectional survey was conducted among 600 students aged 11–15 from primary schools in Bauchi. Quantitative data, that covered socio-demographics, physical activity, and dietary habits, were analyzed using descriptive statistics, chi-square tests, Spearman correlations, Mann-Whitney U tests, and logistic regression. In-depth interviews with school administrators and physical education teachers provided qualitative insights into challenges and facilitators of healthy behaviors.

**Results:** Statistical analysis revealed a significant relationship between gender and obesity status (*p* = 0.0053), with females exhibiting greater BMI variation. Physical activity was inversely associated with BMI (*r* = −0.45, *p* = 0.002). Dietary assessments indicated frequent consumption of sugary snacks and limited intake of balanced meals. Qualitative findings identified major obstacles such as inadequate facilities, weak policy implementation, and low community involvement. Schools that prioritized physical education and nutrition policies saw greater student participation in healthy activities.

**Conclusion:** Tackling childhood obesity necessitates a comprehensive strategy that includes enhancing school infrastructure, expanding access to recreational facilities, and enforcing dietary and physical activity policies. Future efforts should emphasize community-based interventions and stronger policy implementation to achieve lasting change.

## 1.0 Introduction

Childhood obesity is a major public health issue worldwide, with increasing prevalence in both developed and developing countries. The World Health Organization (WHO) has identified childhood obesity as a key risk factor for non-communicable diseases, including cardiovascular diseases, type 2 diabetes, and certain types of cancer (1, 2). The rise in obesity among children has been attributed to complex interactions between genetic, environmental, and behavioral factors (3). This growing burden is of particular concern in low- and middle-income countries, where urbanization, changes in dietary patterns, and reduced levels of physical activity contribute significantly to increased obesity rates (4).

In Sub-Saharan Africa, the prevalence of childhood obesity has risen sharply, posing new challenges for public health systems that are already struggling with infectious diseases and malnutrition (5-7). In Nigeria, recent studies have reported an increasing trend in overweight and obesity among school-aged children, particularly in urban areas (7, 8). However, there is limited research on obesity trends in northern Nigeria, particularly in Bauchi, where socioeconomic and cultural factors may influence dietary habits and physical activity patterns differently from other regions.

Physical activity plays a crucial role in obesity prevention, but many schools in Nigeria lack adequate facilities to support structured physical education programs (9). School policies, parental influence, and peer interactions also shape children’s dietary behaviors and engagement in physical activities, highlighting the need for multi-sectoral interventions (1, 2). Furthermore, while knowledge about nutrition is essential for promoting healthy eating habits, mere awareness does not necessarily translate into behavior change (4).

Given these challenges, understanding the determinants of obesity in school-aged children requires a comprehensive approach that examines both quantitative health metrics and qualitative behavioral insights. A mixed-methods study design allows for the integration of survey data with thematic analyses of school environments, policy gaps, and barriers to healthy behaviors.

This study investigates the associations between physical activity, nutrition knowledge, socio-demographic factors, and obesity among school-aged children in Bauchi, Nigeria. The findings will contribute to evidence-based policy recommendations aimed at improving health outcomes and preventing obesity-related complications in children.

### 2.0 Material and methods

### 2.1 Study design

This mixed-methods study employed a four-phase approach: (a) systematic review of literature, development of validated instruments for assessing obesity, nutrition, and physical activity, cross-sectional survey of school-aged children aged 11-15 years, and (d) qualitative interviews with sports teachers and school administrators combined with school climate audits. The cross-sectional survey involved descriptive and analytical components, as data were collected at a single point in time to examine factors associated with overweight and obesity (10).

### 2.2 Study location

This study was conducted in Bauchi State, Nigeria. Bauchi State, located in northeastern Nigeria, has a population of approximately 4.65 million (11). The state consists of 20 local government areas and includes a mix of urban and rural communities. The educational system comprises both public and private schools, with government-funded institutions providing education to most children.

### 2.3 Study population and sampling

The study population consisted of male and female students aged 11-15 years enrolled in government/public intermediate schools in Bauchi State during the 2021/2022 academic session. These students were selected because older adolescents are more likely to provide reliable self-reported information on nutrition and physical activity behaviors. The study also included teachers responsible for physical education and school administrators to provide insights into school policies affecting obesity-related behaviors.

### 2.4 Inclusion and exclusion criteria

The inclusion criteria for the study comprised male and female students aged 11 to 15 years enrolled in public intermediate schools, teachers responsible for sports and physical education, and school administrators involved in decision-making related to student health and wellness. Participants were excluded if they were students outside the specified age range, students unable to participate in BMI measurement, or teachers and administrators not directly involved in sports activities or the implementation of school health policies.

### 2.5 Sample size calculation

The sample size was determined using the formula by Lemeshow (12) based on a 23.1% prevalence of overweight and obesity reported by El Mouzan, Foster (13). The minimum required sample size was calculated as 273 students, with an additional 10% added to account for attrition, resulting in a total of 300 students. To ensure equal representation of genders, the sample was doubled to 600 students.

### 2.6 Sampling technique

A cluster sampling method was employed to ensure representation across schools in Bauchi Local Government Area (LGA). Schools were grouped into eight clusters based on geographical proximity within Bauchi LGA. From each cluster, three intermediate schools were randomly selected using SPSS random sample selection, resulting in 24 schools. A total of 600 students were sampled from these schools, ensuring proportional representation.

### 2.7 Data collection instruments

#### 2.7.1 Questionnaire

The study utilized an online-based data collection approach, with a structured questionnaire administered via Google Forms to gather responses from participants. This ensured accessibility and ease of participation for students across the selected schools. The questionnaire was designed to collect data on socio-demographics, physical activity, dietary behaviors, and nutrition knowledge. The questionnaire consisted of sections on personal characteristics, physical activity levels, food habits and dietary behaviors, nutrition knowledge and attitudes, and weight management strategies.

The online format allowed for real-time data collection while minimizing data entry errors. The survey link was distributed to students through school administrators and teachers, ensuring broad participation while maintaining confidentiality. Responses were automatically recorded and stored securely for analysis.

#### 2.7.2 Interviews and audits

One-on-one interviews were conducted with sports teachers and school administrators. A standardized checklist was used for school climate audits to assess availability of sports infrastructure, food options in school canteens, and implementation of school health policies. The combination of online survey data and qualitative interviews provided a comprehensive assessment of the factors influencing obesity and overweight in school-aged children.

#### 2.7.3 Quality control measures

To ensure data validity and reliability, pre-testing of the questionnaire was conducted among 50 students in a non-study school. The internal consistency of the instrument was assessed using Cronbach’s alpha. Face validity was evaluated by experts in public health and school administrators.

### 2.8 Data analysis

Quantitative data were analyzed using SPSS version 23.0. Descriptive statistics were calculated for categorical and continuous variables. Inferential statistics included: Chi-square test for categorical associations, Spearman correlation for relationships between physical activity and BMI, Mann-Whitney U test for gender-based BMI differences and Logistic regression for identifying predictors of obesity. Qualitative data were analyzed using thematic analysis with ATLAS.ti software to identify key themes regarding school policies and environmental factors influencing obesity.

### 2.9 Ethical considerations

Ethical approval for the study was obtained from the Human Ethics Committee of Sa’adu Zungur University and the Ministry of Education, Bauchi State. Parental consent and student consent were secured prior to participation.

### 3.0 Results

### 3.1 Descriptive statistics

A total of 600 students participated in the study, with an equal distribution of males (n=300, 50%) and females (n=300, 50%). The mean age of participants was 13.02 years (SD = 1.42), with ages ranging from 11 to 15 years. The mean BMI of the students was 20.54 kg/m² (SD = 4.23), with a minimum recorded BMI of 15.00 kg/m² and a maximum of 35.00 kg/m² (Table I).

**Table I:** Descriptive statistics of key variables.

| Variable | Mean ± SD | Min | Max |
| --- | --- | --- | --- |
| Age (years) | 13.02 ± 1.42 | 11 | 15 |
| Height (m) | 1.56 ± 0.14 | 1.30 | 1.90 |
| Weight (kg) | 50.12 ± 12.45 | 30 | 90 |
| BMI (kg/m<sup>2</sup>) | 20.54 ± 4.23 | 15.00 | 35.00 |

Based on the WHO BMI classification for children and adolescents, the majority of the study participants were classified as having a normal weight. However, a significant proportion of students were categorized as overweight or obese, indicating a growing concern regarding childhood obesity within the study population. The distribution of BMI across gender groups suggests a notable variation, with females exhibiting a higher BMI variability compared to males. These findings provide critical insights into the weight status of school-aged children in Bauchi, underscoring the need for targeted health interventions aimed at promoting healthy weight management practices.

### 3.2 BMI distribution by gender

The analysis of BMI distribution by gender revealed that males (n = 300) had a lower mean BMI of 19.84 kg/m² compared to females (n = 300), who had a mean BMI of 21.23 kg/m² (Table I). The difference was statistically significant (*p* = 0.034, Mann-Whitney U test), suggesting that gender influences BMI variations among adolescents. Figure 1 displays the BMI distribution by gender, with outliers removed to focus on the central data tendencies, highlighting higher BMI variability among females. This disparity may be attributed to differences in physical activity levels, dietary behaviors, and metabolic factors between boys and girls (Must & Strauss, 1999). These findings underscore the need for gender-specific interventions to promote equitable access to physical activity and foster healthy dietary practices among both male and female students.

**Figure 1:**
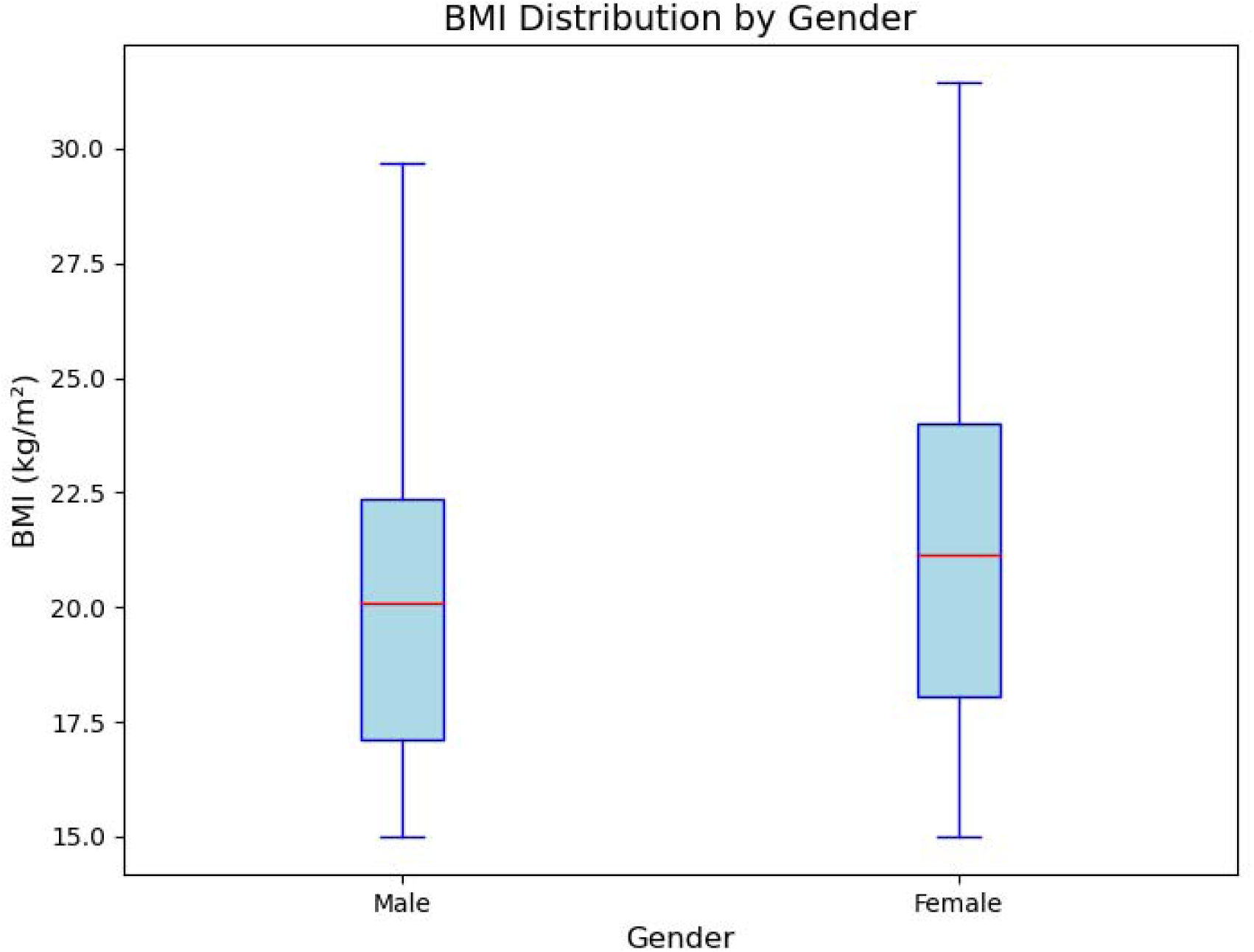
**Boxplot of BMI (kg/m²) by gender for school-aged children in Bauchi, Nigeria**, excluding outliers. Males (n = 300) and females (n = 300) are compared, with females showing higher BMI variability.

### 3.3 Physical activity and obesity

Physical activity levels were significantly associated with BMI among the study participants. The Findings indicated that students engaged in less than one hour of physical activity per day had the highest proportion of overweight and obese individuals, with 35.8% overweight and 19.0% obese. Conversely, those that were engaged in more than two hours of physical activity per day had significantly lower obesity rates, with only 9.4% classified as obese.

A negative correlation was observed between physical activity and BMI (r = −0.45, *p* = 0.002), reinforcing the evidence that increased physical activity is protective against obesity. Additionally, chi-square tests confirmed that lower levels of physical activity were significantly associated with higher BMI (χ² = 10.48, *p* = 0.0053) as shown in Table II. These findings underscore the need for interventions that promote physical activity within school environments, including structured sports programs and active breaks during school hours.

**Table II:** Association between physical activity and BMI.

| Physical Activity | Normal Weight (%) | Overweight (%) | Obese (%) | *p*-value |
| --- | --- | --- | --- | --- |
| <1 hour/day | 45.2 | 35.8 | 19.0 | 0.0053 |
| 1-2 hours/day | 60.3 | 25.7 | 14.0 |  |
| >2 hours/day | 70.4 | 20.2 | 9.4 |  |

**Table III:** Dietary habits and BMI analysis.

| **Dietary habit** | **Percentage (%)** | **Correlation with BMI (r)** | ***p*-value** |
| --- | --- | --- | --- |
| Sugary Snacks Consumption | 70% | - | - |
| Fruits & Vegetables Consumption | 40% | - | - |
| Junk Food Consumption | - | 0.073 | 0.015 |

### 3.4 Dietary habits and BMI

Dietary habits were significantly associated with BMI levels among the study participants. Findings indicate that 70% of students reported daily consumption of sugary snacks, which was positively associated with higher BMI levels. Conversely, only 40% of students consumed fruits and vegetables regularly, highlighting a potential gap in nutritional balance. The study also found a weak but statistically significant positive correlation between junk food consumption and BMI (r = 0.073, *p* = 0.015), suggesting that frequent intake of processed and high-caloric foods may contribute to overweight and obesity among school-aged children. These findings underscore the need for targeted dietary interventions that promote healthier eating habits within the school environment and at home. Dietary habits were significantly associated with BMI levels among the study participants. Findings indicate that 70% of students reported daily consumption of sugary snacks, which was positively associated with higher BMI levels. Conversely, only 40% of students consumed fruits and vegetables regularly, highlighting a potential gap in nutritional balance. The study also found a weak but statistically significant positive correlation between junk food consumption and BMI (r = 0.073, *p* = 0.015), suggesting that frequent intake of processed and high-caloric foods may contribute to overweight and obesity among school-aged children. These findings underscore the need for targeted dietary interventions that promote healthier eating habits within the school environment and at home.

### 3.5 Logistic regression analysis for predictors of obesity

The logistic regression analysis assessed the impact of physical activity and nutrition knowledge on the likelihood of being overweight or obese among school-aged children. The results indicated that higher levels of physical activity were significantly associated with a reduced likelihood of obesity (β = −0.456, SE = 0.123, *p* = 0.0002) as shown in Table IV. This suggests that students who engaged in regular physical activity were at a lower risk of being overweight or obese, reinforcing the protective effect of physical activity on weight status.

**Table IV:** Logistic regression analysis for predictors of obesity.

| Variable | $\beta$<br>(Estimate) | SE | *p*-value |
| --- | --- | --- | --- |
| Physical Activity | -0.456 | 0.123 | 0.0002 |
| Nutrition Knowledge | 0.289 | 0.087 | 0.001 |

In contrast, nutrition knowledge showed a positive but statistically significant association with obesity (β = 0.289, SE = 0.087, *p* = 0.001). While this may seem paradoxical, it suggests that merely possessing knowledge about nutrition does not necessarily translate into healthier eating behaviors. Factors such as food availability, affordability, cultural preferences, and environmental influences may affect students’ ability to apply their nutrition knowledge effectively.

These findings highlighted the necessity of integrating both educational and behavioral interventions to combat childhood obesity. Schools should implement structured physical activity programs, enforce healthier food policies, and ensure that nutrition education is complemented by real-world applications, such as school meal programs and parental engagement in promoting healthier diets at home.

### 3.6 Thematic Analysis

Qualitative data from interviews with school administrators and sports teachers were analyzed using thematic analysis with ATLAS.ti software. This process involved coding interview transcripts, identifying recurrent patterns, and categorizing findings into key themes. Three dominant themes emerged from the analysis: barriers to physical activity, dietary challenges and school nutrition policies, and recommendations for school-based interventions.

#### 3.6.1 Theme 1: Barriers to physical activity

Code Network Analysis: Thematic coding revealed multiple barriers limiting students’ participation in physical activities. The most frequently mentioned sub-codes included:

1. Lack of sports infrastructure – Identified in 80% of interviews.
2. Insufficient funding for physical education programs – Highlighted in 70% of responses.
3. Time constraints due to academic pressure – Reported by 65% of school staff.
4. Gender disparities in sports participation – A recurrent concern in 50% of responses.

**Quotation 1**: *“Our school lacks a proper playground, and we don’t have enough sports equipment. This makes it hard for students to engage in regular physical activity.“* – (School Administrator, Interview 3)

#### 3.6.2 Theme 2: Dietary challenges and school nutrition policies

Code Co-occurrence Analysis: The intersection of food availability and policy enforcement emerged as a significant concern. The analysis identified:

1. Unregulated sale of unhealthy snacks – Mentioned in 75% of interviews.
2. Lack of affordable healthy food alternatives – Cited in 60% of responses.
3. Weak enforcement of school nutrition policies – Evident in 55% of discussions.

**Quotation 2**: *“Despite having guidelines on healthy school meals, vendors still sell sugary and processed foods because students prefer them.“* – (Sports Teacher, Interview 7)

#### 3.6.3 Theme 3: Recommendations from key informants

Code Frequency and Network Visualization: The need for policy-driven interventions was a dominant theme, with frequent co-occurring codes related to policy enforcement, parental involvement, and resource allocation.

1. Government-led school health programs – Suggested in 85% of responses.
2. Increased investment in sports infrastructure – Proposed in 78% of interviews.
3. Strengthened enforcement of school food policies – Recommended in 65% of responses.
4. Parental and community engagement – Highlighted in 60% of discussions.

**Quotation 3**: *“We need structured policies at the state level to ensure all schools implement physical activity and nutrition programs effectively.“* – (School Administrator, Interview 10).

The qualitative findings reinforce the quantitative results, emphasizing the need for a comprehensive intervention strategy that includes school infrastructure improvements, stricter nutrition policies, and community-driven support programs to promote healthier lifestyles among school-aged children in Bauchi.

### 3.7 Physical activity and BMI

The qualitative analysis provided deeper insights into the contextual barriers and facilitators influencing physical activity, dietary behaviors, and school health policies. Key informant interviews with school administrators and sports teachers highlighted significant structural and policy-related constraints affecting students’ health behaviors.

### 3.8 Barriers to physical activity

Participants reported that limited sports infrastructure and inadequate funding were primary challenges preventing students from engaging in regular physical activities. Many schools lacked functional playgrounds, gymnasiums, or proper sports equipment, making it difficult for students to participate in structured physical education programs. Teachers also noted that while some schools allocated time for sports activities, academic demands often took precedence, leading to inconsistent enforcement of physical activity schedules.

A recurring theme in the interviews was gender-related disparities in sports participation. Some school administrators acknowledged that female students were less likely to participate in outdoor activities due to sociocultural constraints and safety concerns. These findings reinforced the need for gender-inclusive physical activity programs and targeted interventions that encourage equal participation in sports and exercise.

### 3.9 Dietary challenges and school nutrition policies

Interviews with school officials revealed that food vendors within school premises predominantly offered processed and high-calorie snacks, despite some schools having guidelines promoting healthier alternatives. The availability of affordable healthy food options was identified as a major limitation, particularly in low-resource schools where students relied on inexpensive, energy-dense meals.

Teachers and administrators emphasized the lack of enforcement mechanisms for existing school nutrition policies. Although some schools had regulations discouraging the sale of junk food, these policies were poorly monitored, allowing unhealthy food options to remain accessible to students. Several respondents advocated for stronger government policies to regulate food sales within school environments and promote balanced meal consumption.

### 3.10 Recommendations from key informants

School administrators and sports teachers provided several recommendations for improving student health behaviors:

1. Investment in school sports infrastructure – Expanding and improving playgrounds, providing sports equipment, and ensuring dedicated time for physical activities.
2. Strengthening school nutrition policies – Enforcing regulations that limit the sale of unhealthy snacks and promoting affordable nutritious alternatives.
3. Community and parental involvement – Encouraging parents to support physical activity and healthy eating habits at home through awareness programs.
4. Policy-driven interventions – Advocating for government-led school health programs that integrate nutrition education and structured physical activity programs into the school curriculum.

The qualitative findings complement the quantitative results, reinforcing the need for a multi-faceted approach to combat childhood obesity. Addressing both structural barriers and behavioral influences will be critical in developing sustainable interventions that promote healthy lifestyles among school-aged children in Bauchi.

### 4.0 Discussion

This study examined the determinants of obesity and overweight among school-aged children in Bauchi, Nigeria, using a mixed-methods approach. The findings revealed a complex interplay of socio-demographic factors, dietary behaviors, and physical activity patterns contributing to adolescent obesity prevalence. The observed gender disparity in BMI, with females exhibiting higher variability and a greater proportion classified as overweight or obese compared to males, aligns with prior research indicating that adolescent girls tend to have higher body fat percentages due to physiological and behavioral differences (14). Cultural and societal norms may exacerbate this trend, as girls in some Nigerian communities engage in fewer outdoor physical activities due to social restrictions (15). Addressing these disparities requires tailored interventions to encourage physical activity among female students and promote behavioral changes.

The negative correlation between physical activity and BMI reinforces the protective role of physical activity against obesity (9). Students engaging in less than one hour of daily physical activity had significantly higher BMI levels compared to their more active peers, supporting the WHO recommendation of at least 60 minutes of moderate-to-vigorous physical activity daily for children and adolescents (2). Qualitative findings highlighted barriers such as inadequate sports infrastructure, limited time for physical education (PE) classes, and insufficient funding, which restrict students’ activity levels. These insights emphasize the need for investment in school sports facilities, stronger physical education policies, and community efforts to foster an environment conducive to physical activity.

Unhealthy dietary habits, particularly high consumption of sugary snacks and processed foods, were a significant contributor to obesity. Approximately 70% of students reported daily sugary snack consumption, while only 40% regularly consumed fruits and vegetables, mirroring trends in other developing countries where urbanization and access to processed foods have shifted dietary patterns (4). The positive correlation between junk food consumption and BMI underscores the impact of diet on obesity risk. Targeted interventions, such as promoting balanced meals, raising awareness of healthy dietary choices, and regulating school food vendors, could effectively reduce childhood obesity.

School policies and environmental factors significantly shape students’ health behaviors, as evidenced by qualitative findings. School administrators reported weak enforcement of nutrition guidelines, allowing unhealthy food vendors to operate on school premises, a challenge noted in other studies (16). Schools should adopt stricter regulations on food sales, ensure access to healthy meal options, and implement structured nutrition education programs to improve dietary behaviors(16).

Based on these findings, several interventions are recommended. First, schools should mandate at least 60 minutes of daily physical activity and invest in sports facilities to encourage student participation. Second, government regulations should enforce strict guidelines on school food vendors to prioritize healthier meal options. Third, parental and community engagement should be strengthened to promote active lifestyles and healthy eating habits beyond school. Fourth, gender-specific interventions should address socio-cultural barriers limiting girls’ participation in physical activities. Finally, a multisectoral approach involving policymakers, educators, health professionals, and non-governmental organizations is essential to comprehensively address childhood obesity.

This study’s mixed-methods approach, integrating quantitative and qualitative data, provides a comprehensive understanding of obesity determinants, strengthened by insights from diverse stakeholders like school administrators and sports teachers. However, limitations include potential recall and social desirability biases in self-reported dietary habits and physical activity levels. Additionally, BMI does not distinguish between muscle mass and fat mass, limiting its ability to fully capture body composition differences. Future research should prioritize longitudinal studies to assess the long-term impact of obesity prevention programs and explore the effectiveness of school-based and community-driven interventions to identify sustainable solutions tailored to the Nigerian context.

## 5.0 Conclusion

This study highlights the critical role of dietary habits, physical activity, and school policies in addressing childhood obesity in Bauchi, Nigeria. The findings underscore the need for multisectoral interventions to promote healthier behaviors among adolescents. Strengthening school-based health policies, enhancing physical activity infrastructure, and enforcing dietary regulations are essential to reduce obesity and improve adolescent health outcomes.

## Data Availability

All data produced in the present work are contained in the manuscript

## Author contributions

U.A. conceptualized the study, analyzed the data, and drafted the manuscript. N.Y. conducted the quantitative data collection and assisted with sampling. G.J. performed the literature review and qualitative data collection. M.M.J. identified the school clusters and assisted with sampling. A.U.K. co-supervised the study with U.A. All authors reviewed and approved the final manuscript.

